# The efficacy of soap against schistosome cercariae: A systematic review

**DOI:** 10.1101/2022.05.02.22274382

**Authors:** Jiaodi Zhang, Ana K. Pitol, Laura Braun, Lucinda Hazell, Michael R. Templeton

## Abstract

**Background:** Schistosomiasis is a parasitic disease that is endemic in 78 countries and affects almost 240 million people worldwide. It has been acknowledged that an integrated approach that goes beyond drug treatment is needed to achieve control and eventual elimination of the disease. Improving hygiene has been encouraged by World Health Assembly, and one aspect of good hygiene is using soap during water-contact activities, such as bathing and doing laundry. This hygiene practice might directly reduce the skin exposure to cercariae at transmission sites. A qualitative systematic review was carried out to investigate the efficacy of soap against schistosome cercariae and to identify the knowledge gaps surrounding this topic.

**Methodology:** Six online databases were searched between 5^th^ and 8^th^ July of 2021. Records returned from these databases were screened to remove duplicates, and the remaining records were classified by reading titles, abstracts, and full-texts to identify the included studies. The results were categorised into two groups based on two different protective mechanisms of soap (namely, damage to cercariae and protection of skin).

**Conclusions:** Limited research has been conducted on the efficacy of soap against schistosome cercariae and only 11 studies met the criteria to be included in this review. The review demonstrates that soap has the potential of protecting people against schistosome cercariae and there are two protective aspects: (1) soap affects cercariae adversely; (2) soap on the skin prevents cercariae from penetrating the skin. Both aspects of protection were influenced by many factors, but the differences in the reported experimental conditions, such as the cercarial endpoint measurement used and the cercaria numbers used per water sample, lead to low comparability between the previous studies. This review demonstrates that more evidence is needed to inform hygiene advice for people living in schistosomiasis endemic areas.

**Author summary:** Schistosomiasis affects millions of people living in low- and middle-income countries lacking safe access to water, sanitation and hygiene (WASH). Schistosomiasis is mainly controlled by drug treatment with praziquantel, but people become infected again if they continue to come into contact with cercaria-contaminated water. Despite all the efforts that have been made, this disease remains a major public health problem in disease-endemic regions. The use of soap during water contact, as part of good hygiene practice, may be an effective complementary control method as it might reduce the penetration of schistosome cercariae into the skin, thereby reducing the likelihood of reinfection. We conducted a systematic review to summarise previous research into the efficacy of soap against schistosome cercariae, and to identify current knowledge gaps to inform future studies. The information gathered from this review can guide hygiene advice regarding soap use in endemic areas to reduce the spread of schistosomiasis.

## Introduction

Schistosomiasis is a parasitic disease caused by trematode worms of the genus *Schistosoma* that affects approximately 240 million people worldwide [1]. It is estimated that the global burden of schistosomiasis is 1.6 million disability-adjusted life years (DALYs) according to the Global Burden of Disease Study 2019 [2]. This disease occurs because of skin-contact with water contaminated by cercariae, which are a larval stage of the schistosome parasite. The life cycle of the parasite involves the excretion of *Schistosoma* eggs into freshwater via the faeces or urine of infected individuals. The eggs then hatch into miracidia which infect the intermediate hosts, water snails. The snails release cercariae, which can penetrate human skin, develop into adult worms and produce eggs inside humans.

Several interventions can be adopted to control schistosomiasis transmission, such as preventive chemotherapy and snail control. Preventive chemotherapy, namely mass drug administration with praziquantel, has been the primary control measure to date since it is cost-effective and relatively safe [1,3,4]. However, this intervention cannot kill juvenile worms nor prevent re-infection which is likely in communities relying on contaminated freshwater for daily water activities. The mass drug administration also relies heavily on the coverage level of drug distribution and ongoing financial support [5,6]. Snail control using molluscicides has also proven to be an effective way to interrupt disease transmission and reduce disease prevalence [7–9]. Nevertheless, there are concerns that molluscicidal chemicals might adversely affect aquatic lifeforms, and some studies have shown that mollusciciding is labour-intensive and difficult to apply effectively in some settings [10].

Despite the efforts and progress made, schistosomiasis remains a major public health problem in disease-endemic regions. In the new road map for neglected tropical diseases, the World Health Organisation (WHO) has proposed a target of eliminating schistosomiasis as a public health problem by 2030, aiming to reduce the proportion of heavy intensity infections to less than 1% [11]. In the World Health Assembly (WHA) resolution 65.21, water, sanitation and hygiene (WASH) interventions are encouraged as components of schistosomiasis control and elimination in disease-endemic areas [12], emphasising the importance of WASH as part of a multi-faceted and sustainable disease control strategy [13,14].

Hygiene can be defined as personal and household-level practices (e.g., handwashing, bathing, laundry) which are aimed at achieving cleanliness, maintaining health and preventing disease [6,11]. One aspect of good hygiene is the use of soap when washing, and this hygienic practice is important to prevent many infectious diseases [15,16], such as diarrhea caused by enteric pathogens [17], trachoma [18] and acute respiratory diseases [19]. Apart from soap, alternative handwashing materials such as ash can be used. However, it remains unclear whether the use of ash during handwashing is effective at protecting people from viral and bacterial infections [17, 23].

The use of soap might play a vital role in controlling schistosomiasis and Grimes *et al*. have suggested that this role might have two aspects [21]. The first aspect is that people who are using soap during water contact may be protected from infection. The second aspect is that soap may kill other schistosome life stages in water, including miracidia, cercariae and snails, and thereby help prevent other people from becoming infected. Since infection only occurs when cercariae penetrate human skin during water-contact activities and several of these common water-contact activities such as laundry, bathing and handwashing involve the use of soap, it is crucial to understand the interaction between soap use and cercaria infectivity.

Some field studies have been carried out in disease-endemic regions to investigate the effectiveness of soap use in preventing schistosomiasis, but the results remain inconclusive, since some observed a positive effect and some did not. Endod (*Phytolacca dodecandra*) berries are a typical local soap in Ethiopia [22]. These small berries can be used to wash clothes after being dried, grounded and placed in water [23]. One field study in Ethiopia reported a significant reduction in the prevalence of female infection after soap containing ground Endod berries were distributed to people washing clothes, though the prevalence in males increased [24]. This might be because the majority of washing activities are likely to be performed by women and girls [21]. The result of another study suggested the possible positive influence of soap, as this study found no significant association between clothes-washing and schistosomiasis infection, with a high reported soap-use rate of 98.4% during laundry in the rural communities of Zanzibar [25]. However, there are other studies which did not show that soap has any positive protective effect against schistosomiasis. A field study in Niger found that the use of soap did not prevent children from being infected, though almost all women claimed to use soap when washing their children at the transmission sites [26]. Also, no significant relationship was found between schistosomiasis infection and the use of handwashing facilities equipped with available soap and water [27,28]. These conflicting results may be due to the fact that the protective effect of soap is highly human behaviour-specific [21], in terms of the different practices of using the soap, and the disease prevalence can also be influenced by many non-hygiene related factors. Therefore, results obtained from controlled experiments are likely to be more informative about the efficacy of soap against schistosome cercariae than observational field studies.

The increased use of soap has the potential to be adopted as an effective intervention to prevent schistosomiasis. This research aims to summarise the knowledge obtained from previous experiments in terms of soap use against schistosome cercariae and identify knowledge gaps. The information gathered from this research will help to optimise soap use from a disease prevention viewpoint and form the basis of hygiene practice guidelines (e.g., soap types, amounts, exposure times) to control the spread of schistosomiasis.

## Methods

Our systematic review followed the guidelines of the Preferred Reporting Items for Systematic reviews and Meta-Analyses (PRISMA). This review was not registered, and the protocol was not prepared. Papers were searched between 5^th^ and 8^th^ July of 2021 in six databases: Web of Science, Scopus, PubMed, The British Library, Google Scholar, and China National Knowledge Infrastructure (CNKI). All languages and document types were included, and databases were searched from inception to the present day. All databases were searched for any combination of common schistosomiasis-related terms (*Schistosom*\*, cercaria*, *mansoni, haematobium, japonicum*, Bilharz*, snail fever) and hygiene-related terms (soap, detergent, endod, *Phytolacca dodecandra*, ash, laund*, bath*, shower*, handwash*, clean*, wash*, hygien*) in the title. A full list of search terms and search strategies for each database can be found in S1 Supporting Information.

### Classification criteria

A classification system was developed to screen and classify the records (JZ). First, duplicate records were removed and assigned the code 1. Remaining records were classified by screening titles, and when the titles were not relevant to the scope of this review, the records were removed and assigned the code 2. Afterwards, the records were classified by reading their abstracts. Records were excluded and assigned the codes 3-5 when their abstracts showed the studies were (1) not experiment-based studies (2) not about the use of soap (3) not about the effect of soap against cercariae. The full texts of the remaining records were read. When the papers met the same criteria for screening abstracts, those papers were assigned the codes 6-8. Chapter titles of books were screened; however no relevant studies were found. The remaining papers were included in this review and assigned the codes 9-10 based on the two protective aspects of soap. Additional studies which were referenced in the already included papers in a way indicating that these studies met the objective of this review were also retrieved for the full texts to be read, included and assigned the corresponding codes. A flow diagram (Fig 1) were adapted from the PRISMA 2020 flow diagram with minor changes to summarise the classification process. The classification results including the classification process, codes and a list of the excluded and included records are summarised in the S1 Table.

**Fig 1.**
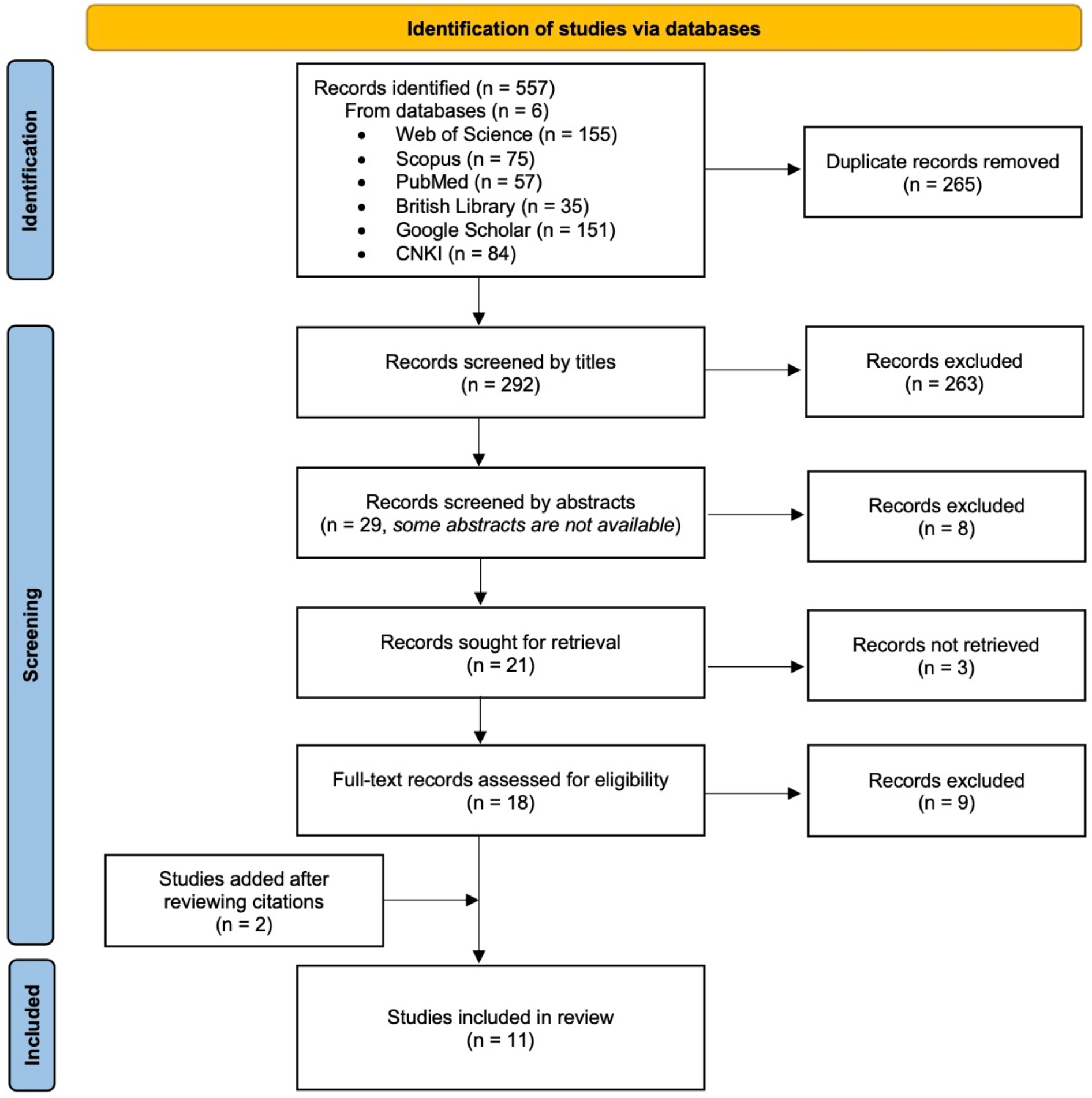
PRISMA flow diagram.

Full texts were acquired from Imperial College London Library and the British Library. Non-English papers were translated using Google Translate first to decide whether the study should be included. Then, translations of those included papers were checked by native speakers.

### Data extraction

Two tables (S1 Table) were developed based on two aspects of potential protection from soap: an adverse effect of soap on cercariae, and a reduction of cercariae penetration into the skin. The first two authors (JZ and AKP) independently extracted data on potential protection from soap (S1 Table). Any discrepancies in the extracted data between the two authors were discussed and resolved.

Some washing powders and detergents were tested in the included papers. These are described as powder soap in the sections below.

### Synthesis of results and data analysis

Parts per million (ppm) is used to compare the soap concentrations in the sections below, since the majority of soap concentrations in included papers were presented in ppm. Other concentrations in mg/L or mol/L were converted to ppm based on the assumption that the liquid density is 1 g/mL. Some other assumptions were made to include the data (see caption of Fig 2).

**Fig 2.**
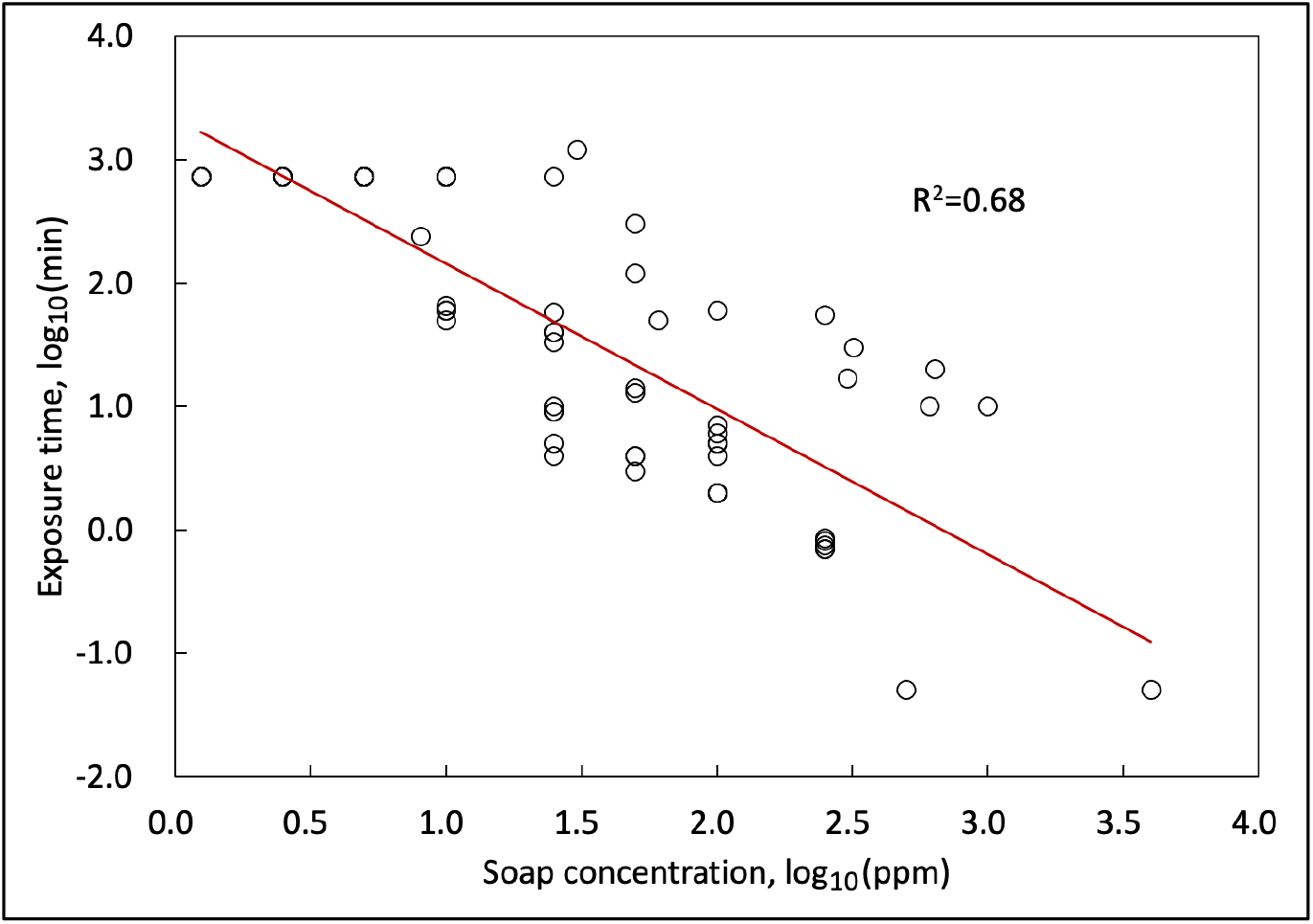
Soap concentrations and exposure times required to achieve 100% cercariae mortality. [23,37,42,44,45]. Data are generated from soaps which were tested in six studies. Several assumptions were made as follows: (1) The study by Mimpfoundi & Dupouny (1983) stated that the cercariae survived more than 12 hours; therefore, it was assumed that cercariae only survived 12 hours in order to include this research [44]. (2) LC90 was used as the dose for 100% mortality in the study by Monkiedje, Anderson & Englande (1991) [42]. (3) Only one individual compound existed in both soaps tested in the study by Pacheco & Jansen (1951) [45]. (4) In the studies by Mimpfoundi & Dupouny (1983) and Okwuosa & Osuala (1993), the exposure time of “immediate cercaricidal effect/death” was assumed to be 3 seconds based on our estimation that at least 3 seconds are required to determine cercariae death [37,44]. (5) The concentrations in mg/L and mol/L were converted to ppm, assuming that the liquid density is 1g/mL.

**Fig 3.**
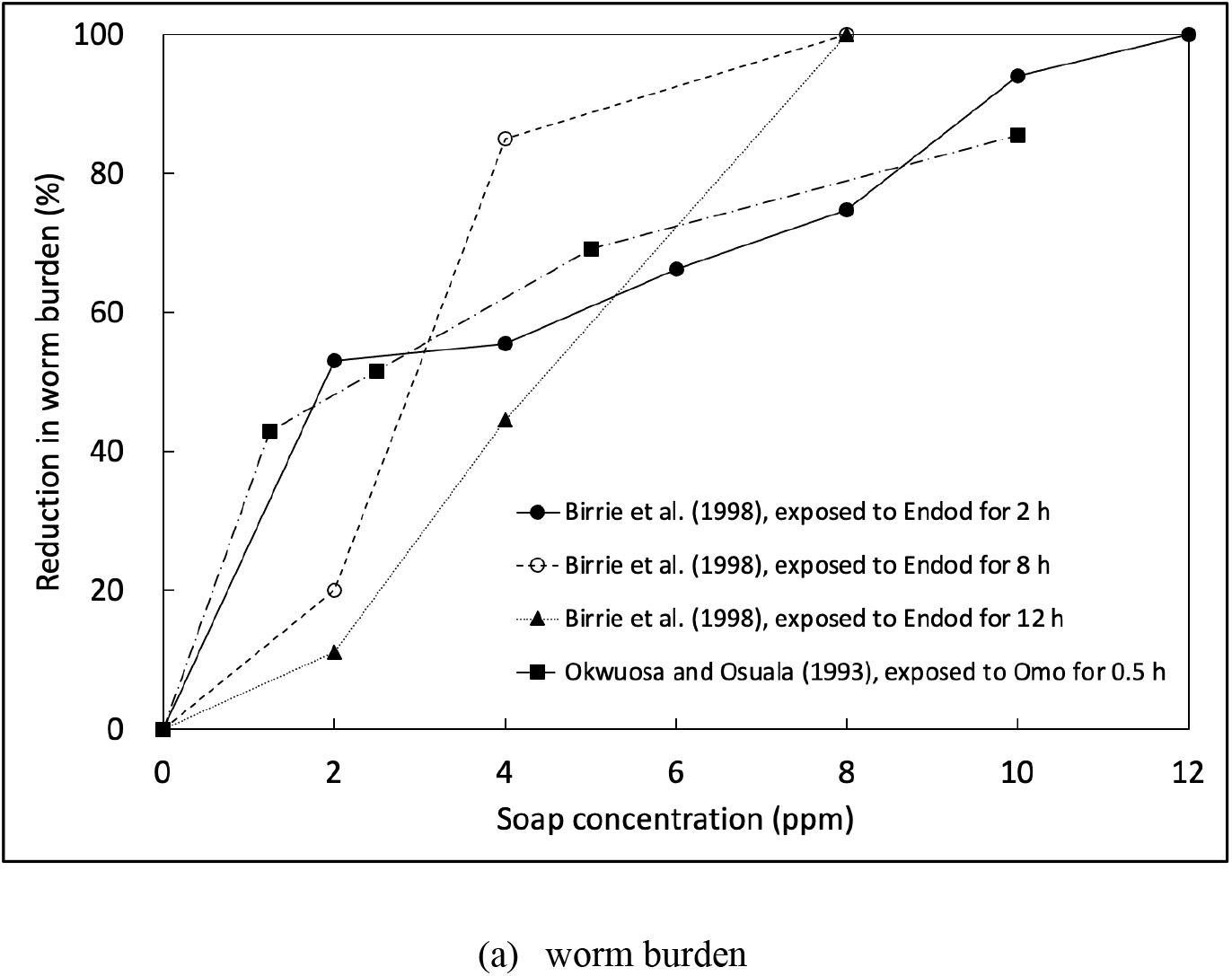
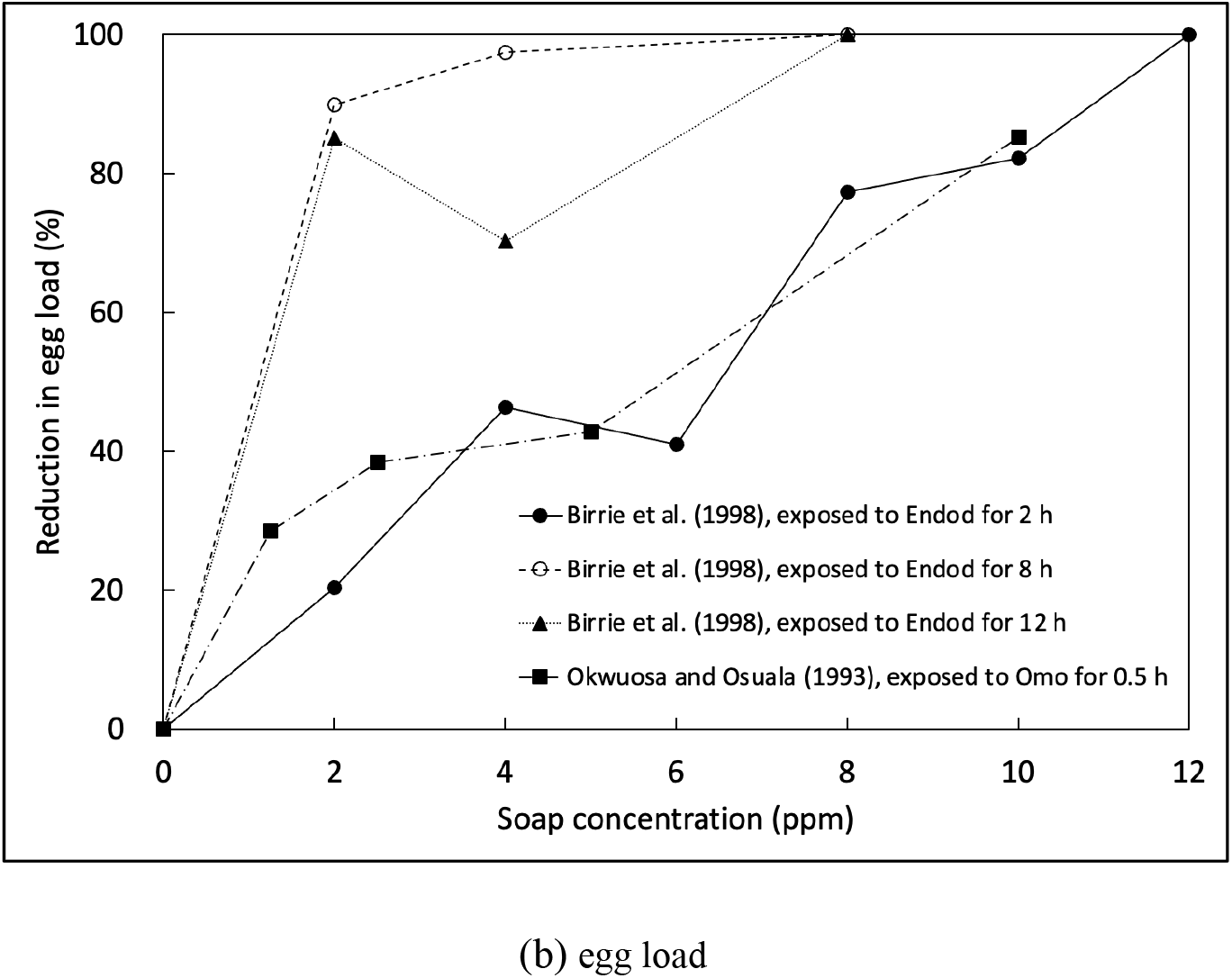
The reduction in worm burden (a) and egg load (b) under different soap treatment conditions. [32,37]. The reduction in worm burden was calculated by comparing the average of worms recovered in the soap-treated group with that in the untreated group.

Regarding the soap concentration in ppm during handwashing, a rough estimate was performed as below. One study has found that it takes 0.4 ∙9 mL of liquid soap for each handwash [29]. According to the United Nations Children’s Fund, the volume of water consumed can be up to 0.3 - 2 L per handwash, depending on whether or not the water is running when scrubbing the hands with soap[30]. Given the data above, it is estimated that the average soap concentration during handwashing can range from 200 ppm (0.4 mL / 2 L) to 30000 ppm (9 mL / 0.3 L).

The results of the two soap protective aspects were presented in different ways. For the effect of soap on cercariae, different measures (e.g., time taken to kill all cercariae at a concentration, worm burden, egg load) were used depending on the cercaria endpoint assessed and one model simulation for the effect on cercaria mortality was performed using SPSS software (Version 23, IBM). For the soap protection on skin against cercaria penetration, the protection which is the reduction in worms recovered from soap-treated mice/rats compared with the control was calculated to show the results. All the figures except Fig 1 were drawn using the Microsoft Excel (Version 2019).

## Results and discussion

The result of our classification process is shown in the flow diagram (Fig 1). A total of 557 records were returned and 292 records remained after duplicates were removed. After screening titles and abstracts, 21 records were sought for full-text retrieval including the records whose abstracts were not available and two books, while three full-texts were not available. All the (sub)chapter titles of those two books were read and the full texts of the (sub)chapters were read when the titles showed they were relevant to the review, though no relevant studies in those books were found. Two additional studies which were referenced in already-included studies were also added to the review. In the end, 11 studies were included in the analysis of this systematic review.

### Soap type

Three major types of soap were investigated in the included studies: locally produced soap, commercial soap, and experimental soap (Table 1). Endod, a locally produced soap, was not only toxic to cercariae but also to miracidia, snails and fish [31–33]. Its molluscicidal activity has received extensive attention since this plant-derived molluscicide is argued to be more environmentally friendly and economical than synthetic molluscicides [24,33,34]. The protective effect of some commercial soaps (e.g., Dettol, Lux, Omo) was also explored in other studies [35–37]. In addition, two kinds of experimental soap formulations, Schistopel and soaps containing essential oils, were tested. Schistopel is an experimental soap formulation with hexachlorophene as a bacteriostatic agent [38], while other experimental soaps incorporated essential oils extracted from local plants into soap formulations [36,39]. Similar to endod, some essential oils showed promising toxicity against the intermediate hosts of *Schistosoma* spp. and could be potentially used as a molluscicide [40].

**Table 1.** Summary of the soaps considered in the included papers.

| Soap type | Soap name | # of studies | Protective effect | Cercarial endpoint |
| --- | --- | --- | --- | --- |
| Local soap | Endod | 3 | Effect on cercariae | Mortality<sup>a</sup>, motility<sup>b</sup> |
|  |  |  | Protection on skin | Viability<sup>c</sup> |
|  | Local soap in Nigeria | 1 | Effect on cercariae | Mortality |
| Commercial soap | Omo | 2 | Effect on cercariae | Mortality, viability |
|  | Elephant | 1 | Effect on cercariae | Mortality |
|  | Surf | 1 |  |  |
|  | Action | 1 |  |  |
|  | Flash | 1 |  |  |
|  | Pax | 1 |  |  |
|  | Genie | 1 |  |  |
|  | Sol | 1 |  |  |
|  | Eucalipto | 1 | Protection on skin | Infectivity<sup>d</sup>, viability |
|  | Lux | 1 |  |  |
|  | Dettol | 1 | Effect on cercariae | Mortality, motility, morphology<sup>e</sup> |
|  |  | 1 | Protection on skin | Viability |
| Experimental soap | Schistopel | 2 | Protection on skin | Infectivity, viability |
|  | Soaps containing essential oils | 2 | Protection on skin | Infectivity, viability |
| Others | Sodium oleate | 1 | Effect on cercariae | Mortality |
|  | Sodium ricinoleate | 1 |  |  |

<sup>a</sup>Mortality = Cercaria's survival percentage
<sup>b</sup>Motility = Cercaria's ability to move
<sup>c</sup>Viability = Cercaria's ability to penetrate the skin and develop into schistosomes [41]
<sup>d</sup>Infectivity = Cercaria's ability to attach to or penetrate skin [41]
<sup>e</sup>Morphology = Cercaria's shape

### Protective roles against cercariae

Based on the findings from the included studies in this review, the role of soap against schistosome cercariae has two aspects: (1) soap has adverse effects on cercariae; (2) soap gives protection against cercariae penetration into the skin. Different adverse endpoints of cercariae were assessed including mortality, motility, morphological change, infectivity and viability. For the first three endpoints, microscopes were the tool for observing cercariae. For infectivity and viability, mice/rats were exposed to cercariae by immersing their tails in water and the worm burden or other indices were determined after exposure.

### Protective role 1: the adverse effect of soap on cercariae

#### The adverse effect on cercaria mortality

The results suggest that soap is less toxic to schistosome cercariae than other chemicals, such as niclosamide and chlorine [42,43]. For example, the 50% lethal dose of niclosamide when treating cercariae for 4 hours was 3650 times lower than that associated with endod [42]. Commercial soap tended to be more toxic than local soap since the commercial ones needed less time to kill cercariae than the local Nigerian soap at the same concentration [37]. Endod, also showed a less lethal effect than commercial soaps in two other studies [23,37,44].

As shown in Fig 2, the lethal effect of soap was associated with soap concentration and exposure time, and the log-transformed data can fit well to a linear regression model with a reasonable R^2^ of 0.68. The exposure time required to achieve 100 % cercarial mortality is reduced with increasing soap concentration [23,37,44,45]. The soap’s toxicity might be also affected by other factors, such as water hardness; one study found that an increase in water hardness increased the cercaricidal effect of powder soaps [44].

#### The adverse effect of soap on cercaria motility and morphological change

The motility and morphology of cercariae can also be affected by exposure to soap. Pacheco and Jansen presented cercaria motility in three different phases including sinking to the bottom, slower movement and immobilization which was also used to determine death [45]. Even though some cercariae were not killed by soap, their swimming behaviours became less active [32,35] and this behavioural change was also associated with the concentration and exposure time [32]. Regarding cercariae morphology, only Edungbola (1980) reported significant morphological damage, including evaginated ventral suckers and irreversible severed tails from the body at the 10^−2^ dilution of Dettol which should mean that adding 1 part of Dettol to 99 parts of water [35].

### The adverse effect of soap on cercaria viability

In studies examining the effect of soap on cercarial viability, worm burden and egg load were determined. Worm burden (load) and egg load refer to the worms and eggs recovered from hosts after exposure to cercariae [32,37]. Other indices were also measured in one study, including eggs in faeces of mice, anaemia given by haemoglobin concentration (g/100 ml) and packed cell volume (% age), and hepatosplenomegaly [37].

Similar to was found in studies using cercariae mortality, where the impact of soap increased with soap concentration, both studies have demonstrated that the worm burden reduced with an increase in soap concentration (Fig 4 (a)). Okwuosa and Osuala (1993) found Omo, a commercial powder soap, reduced about 85% of the worm burden and egg load at 10 ppm after treating for 30 minutes, compared to the untreated group [37]. Birrie *et al*. (1998) tested endod and this soap totally suppressed the cercarial viability at 12 ppm after treating for 2 hours, and the worm burden and egg load were also significantly reduced at lower concentrations (Fig 4) [32]. The result of this study also demonstrated that a lower soap concentration was needed to render all cercariae incapable of developing into adult worms at longer soap exposure times, indicating that soap use duration might also play an important role in the ability of soap to affect the viability of cercariae.

### Protective role 2: the protective effect of soap on rodent tails against schistosome cercariae

Another protective aspect of soap against schistosome cercariae is that soap could provide human skin with protection, preventing cercariae from penetrating. In previous studies, living mice or rats were used as the host and soap was applied on their tails before being exposed to cercariae. Cercariae infectivity and viability were the adverse endpoints assessed, as the number of cercariae that failed to penetrate was counted in some studies [36,39,46], and worms were recovered after exposure in all studies [35,36,38,39,46]. Three studies considered the influence of rinsing which could potentially wash off soap; the tails were rinsed with water between soap application and exposure to cercariae [36,38,46]. All these studies demonstrated that soap has the potential of protecting rodent tails against cercariae, and the soap protection can be represented by the reduction in worm burden in comparison with the control. As shown in Table 2, the protection varied among different soaps, which might be due to several influencing factors, such as soap concentration, the times of soap application, rinsing the tails, the rinse duration, and the time interval between soap application, tail rinse and tail exposure to cercariae.

**Table 2.**
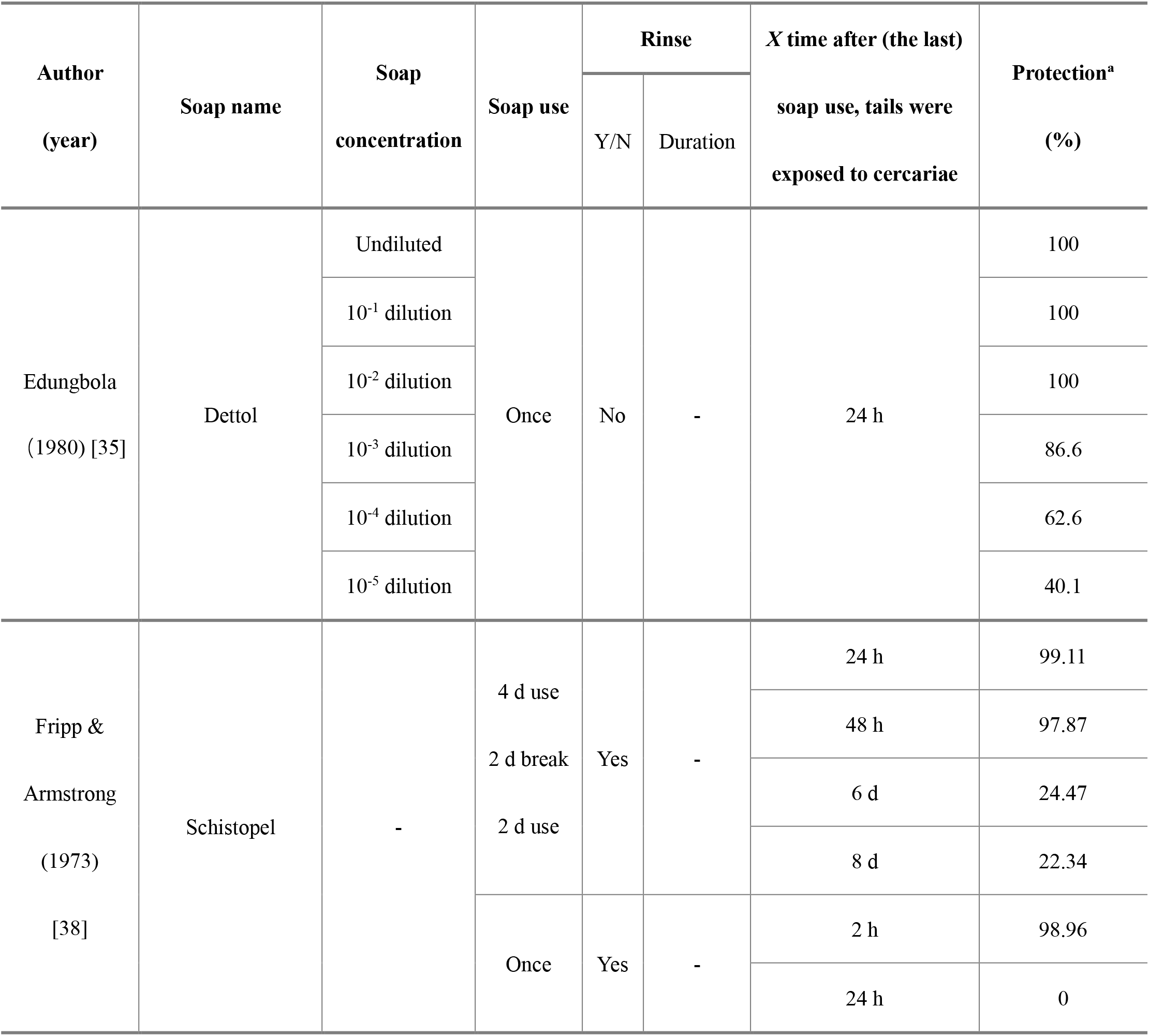
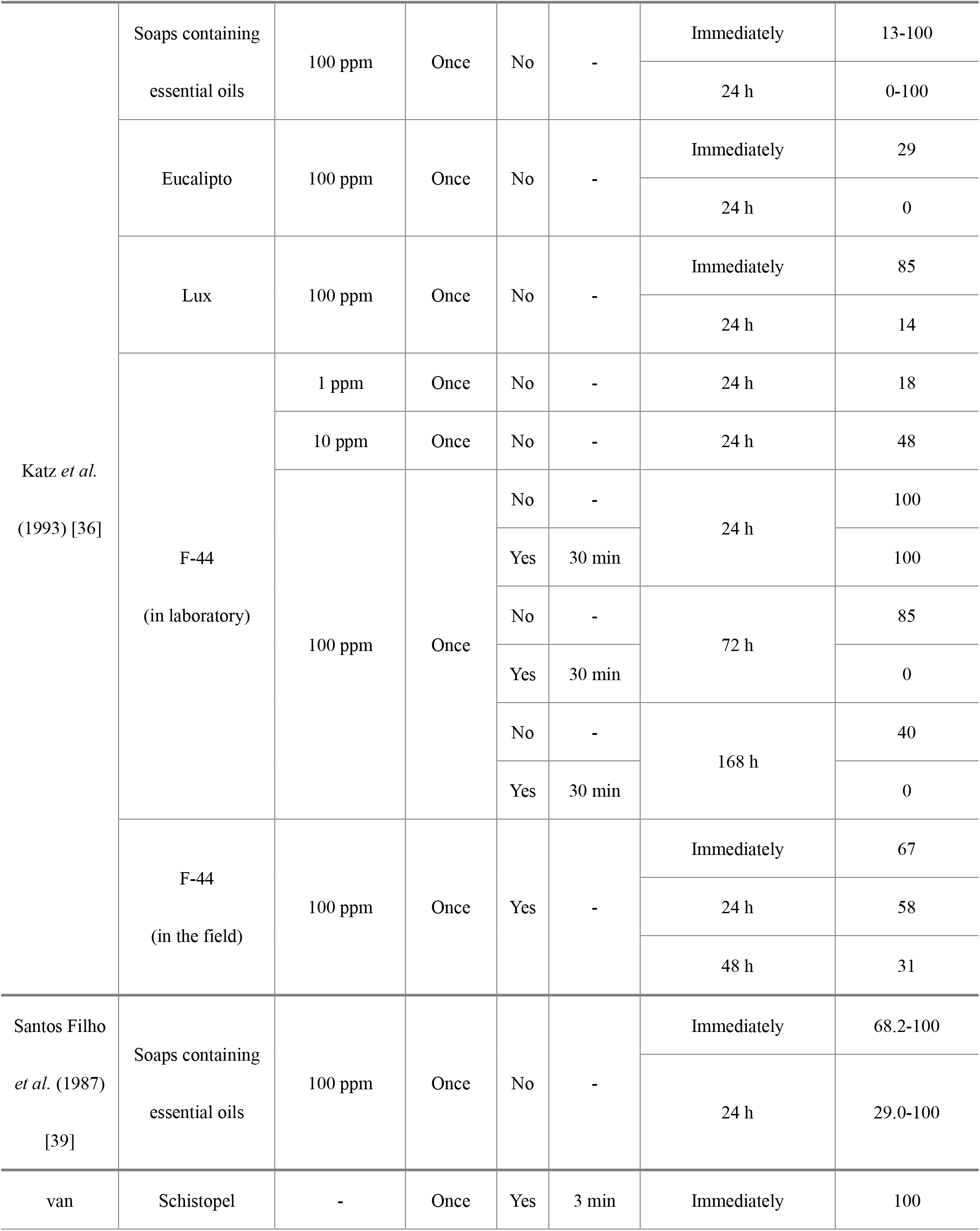
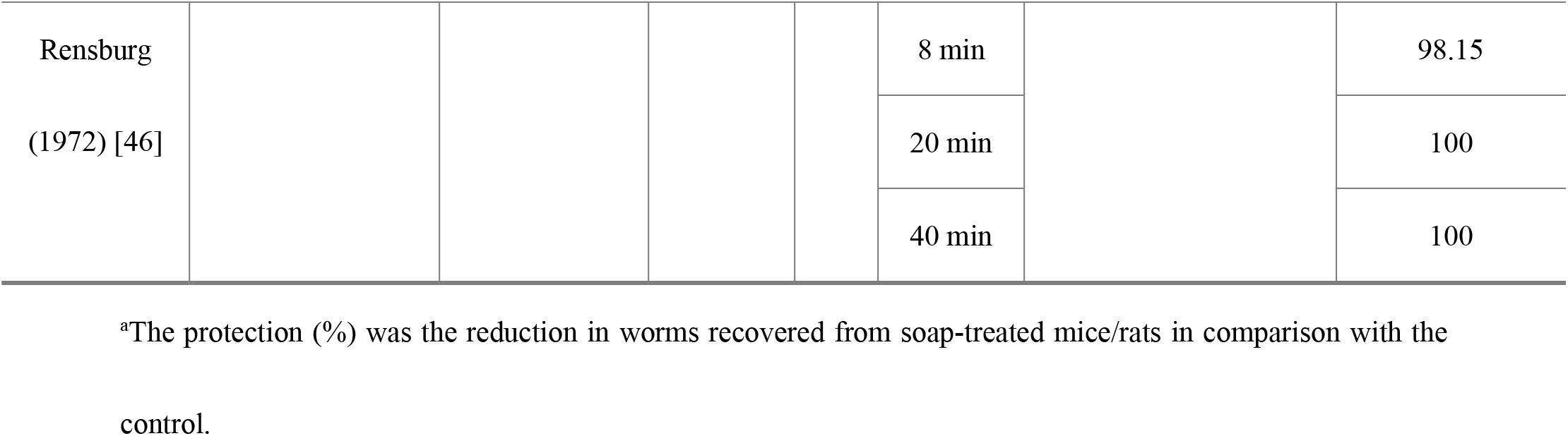
The protective effect of soap on rodent tails under different conditions.

Fripp and Armstrong investigated the protection from an experimental soap called Schistopel under different treatments [38]. They found that 2 hours after using this soap once, this soap provided rodent tails with almost 100% protection when exposed to cercariae for 20 minutes after the tail rinse, but this protection no longer existed when exposed to cercariae 24 hours after using soap [38], highlighting the influence of the time interval between washing and exposure to cercariae. Also, this soap was likely to have an accumulated protection since almost 100% protection was provided if using Schistopel on tails for 6 days and then exposing the tails to cercariae 48 hours after the last wash [38]. This accumulative protective effect might be caused by the bacteriostatic agent in Schistopel, namely hexachlorophene, being absorbed by the skin tissues and not being removed immediately when washing [38]. Another study investigated the effect of rinse duration on Schistopel protection and found that this soap was able to provide protection even after rinsing for 40 minutes [46].

Another two studies investigated the protective effect of soaps containing essential oils [36,39]. In those studies, the essential oil was extracted from fruits of the plant *Pterodon pubescens* and then incorporated into different soap formulations. Both studies found that these soaps protected mouse tails, but the protection varied among different soap formulations, ranging from 13 – 100% when tails were exposed to cercariae immediately after soap application without rinsing the tails. This protection on tails was also inversely related to the time interval between soap application and exposure to cercariae. Among different soaps containing essential oils, F-44 showed the highest protection – 100% even when mouse tails were washed for 30 minutes and exposed to cercariae 24 hours after soap application [36]. Its protection on tails increased with the increase in soap concentration, which was also found with the Dettol soap used by Edungbola (1980), though tails were not rinsed in this study [35]. Also, no protection was obtained from F-44 when exposed to cercariae 72 h or 168 h after rinsing, but partial protection existed when tails were not washed [36], suggesting an influence of rinsing on reduced protection by soap.

Katz *et al*. (1993) also compared the efficacy of experimental soaps containing essential oils and two commercial soaps (Eucalipto and Lux) at reducing cercariae penetration and worm burden [36]. The results suggested that these two commercial soaps had less protective effect compared to those soaps containing essential oils. Eucalipto and Lux gave protection of 29% and 85% to tails immediately after soap application without rinsing, respectively, and the protection no longer existed for Eucalipto and dropped to only 14% for Lux 24 hours after applying soap [36]. However, Edungbola (1980) found that the Dettol soap at the dilution of 10^−2^ provided total protection under the same time interval [35].

## 4 Conclusions

This systematic review demonstrates that soap has the potential to harm schistosome cercariae and provide skin with protection, thereby reducing the likelihood of infection during water-contact activities. Different adverse endpoints of cercariae were used in previous studies (i.e., mortality, motility, morphology, infectivity, viability), and several types of soap were tested (e.g., locally produced soap, commercial soap, experimental soap, etc.).

Soap’s toxicity to cercariae was associated with soap concentration and soap exposure time. However, for the studies on cercariae mortality, most studies did not mention how they evaluated the cercaria death and the number of cercariae used in these studies ranged from 10 to more than 300, which may lead to the differences in the reported results. Compared with the experiments that investigated the concentrations required to kill cercariae in water, lower soap doses were needed to make cercariae lose their capability of penetrating and developing into adult worms.

The protection provided by soap on the skin (represented by rodent tails) has also been revealed by several studies. However, it should be noted that the rodent tails were not rinsed in some experiments, which does not comply with the real-world scenario where rinsing is an essential step involved in almost every soap-related activity. High protection after rinsing was only found in two experimental soaps rather than commercial soaps, and no study investigated if protection on skin from commercial soaps could exist after rinsing, suggesting the importance of future research in this area.

The two protective aspects of soap both varied among different studies, which might be due to different cercarial endpoints assessed, soap ingredients, and experimental conditions (e.g., water characteristics, cercarial number, cercarial age, etc.). For skin protection, there are more influencing factors, such as the rinse duration, the skin-soap contact time, the amount of soap applied, the skin surface area exposed, and the time interval. However, most of the factors mentioned above have not been systematically studied and need further investigation.

Most studies in this review tested *S. mansoni*, and only two studies used *S. haematobium* and *S. mattheei*. Also, there are no studies using more than one species. Therefore, the sensitivity of different species of cercariae to soap remains unclear. In addition, only one study was carried out in the natural environment while others were conducted in the laboratory using cercariae shed from lab-cultivated snails. Future studies need to conduct experiments using both cercariae produced by “wild” snails and water samples collected from disease-endemic areas with varying water qualities.

Overall, there is insufficient knowledge about the impact of soap use against schistosome cercariae, which makes it difficult to propose reliable hygiene guidelines for preventing schistosomiasis, such as guidance on which soap types, amounts, and exposure times will be most protective. With the increasing awareness of the important role of WASH in schistosomiasis prevention, further research is needed to optimise the benefit of soap use and help to incorporate this intervention into multi-faceted schistosomiasis control strategies.

## Supporting information

S1 Supporting Information. Search methodology.

S2 Supporting Information. PRISMA 2020 checklist.

S3 Supporting Information. PRISMA 2020 for abstracts checklist.

S1 Table. Search results.

## Data Availability

All relevant data are within the manuscript and its Supporting Information files.

## Supporting information

**S1 Supporting Information. Search methodology**. Search terms and strategies of each database.

(DOCX)

**S2 Supporting Information. PRISMA 2020 checklist**.

(DOCX)

**S3 Supporting Information. PRISMA 2020 for abstracts checklist**.

(DOCX)

**S1 Table. Search results**. Classification process and codes, a list of excluded and included studies, experimental setups and extracted data.

(XLSX)

## Acknowledgements

We are thankful to the librarians at Imperial College London and the British Library for their help in retrieving the full texts of many studies used in this review.

## Author Contributions

**Conceptualization:** Jiaodi Zhang, Ana K. Pitol, Michael R. Templeton.

**Data curation:** Jiaodi Zhang, Ana K. Pitol.

**Formal analysis:** Jiaodi Zhang.

**Funding acquisition:** Michael R. Templeton.

**Investigation:** Jiaodi Zhang, Ana K. Pitol.

**Methodology:** Jiaodi Zhang, Ana K. Pitol, Laura Braun, Lucinda Hazell, Michael R. Templeton.

**Supervision:** Michael R. Templeton.

**Visualisation:** Jiaodi Zhang, Ana K. Pitol.

**Writing – original draft:** Jiaodi Zhang.

**Writing – review & editing:** Jiaodi Zhang, Ana K. Pitol, Laura Braun and Michael R. Templeton.

