## Supplementary material for "The efficacy of soap against schistosome cercariae: A systematic review": S1 Supporting Information. Search methodology.

**S1 Supporting Information. Search methodology.** Search terms and strategies of each database

**1 Search terms**

**1.1 Schistosomiasis terms**

- Disease name: *Schistosom**, *Bilharz**, *Snail fever*
- Cercariae-related: *(Schistosom*)*, *Cercari**, *mansoni*, *haematobium*, *japonicum*

**1.2 Soap terms**

- Soap: *Soap**, *Detergent*, *Endod*, *Phytolacca dodecandra*, *Ash*
- Water-contact activities: *Laund**, *Bath**, *Shower**, *Handwash**, *Clean**, *Wash**
- Hygiene: *Hygien**

**2 Search strategy**

- **Web of Science: Advanced search, all databases (Web of Science Core Collection, BIOSIS Citation Index, CABI: CAB Abstracts®, KCI-Korean Journal Database, MEDLINE®, Russian Science Citation Index, SciELO Citation Index)**

TI=(Schistosom* OR Cercari* OR mansoni OR haematobium OR japonicum OR Bilharz* OR "Snail fever") AND TI=(Soap* OR Detergent* OR Endod OR "Phytolacca dodecandra" OR Ash OR Laund* OR Bath* OR Shower* OR Handwash* OR Clean* OR Wash* OR Hygien*)

- **Scopus: Advanced search**

TITLE(Schistosom* OR Cercari* OR mansoni OR haematobium OR japonicum OR Bilharz* OR "Snail fever") AND TITLE(Soap* OR Detergent* OR Endod OR "Phytolacca dodecandra" OR Ash OR Laund* OR Bath* OR Shower* OR Handwash* OR Clean* OR Wash* OR Hygien*)

- **PubMed: Advanced search**

(Schistosom*[Title] OR Cercari*[Title] OR mansoni[Title] OR haematobium[Title] OR japonicum[Title] OR Bilharz*[Title] OR "Snail fever"[Title]) AND (Soap*[Title] OR Detergent*[Title] OR Endod[Title] OR "Phytolacca dodecandra"[Title] OR Ash[Title] OR Laund*[Title] OR Bath*[Title] OR Shower*[Title] OR Handwash*[Title] OR Clean*[Title] OR Wash*[Title] OR Hygien*[Title])

- **British Library: Main catalogue - Advanced search**

Main title - contains: schistosom* OR cercari* OR mansoni OR haematobium OR japonicum OR bilharz* OR "snail fever"

AND

Main title – contains: soap* OR detergent* OR endod OR "phytolacca dodecandra" OR ash OR laund* OR bath* OR shower* OR handwash* OR clean* OR wash* OR hygien*

- **Google Scholar: Advanced search, where my words occur: in the title of the article**

With all of the words (insert one word in each search): soap, soaps, soapy, soapberry, soapberries, detergent, detergents, endod, "phytolacca dodecandra", ash, laundry, launder, laundering, bath, bathing, shower, showering, handwash, handwashing, clean, cleaning, cleanser, cleansers, wash, washing, hygiene, hygienic

With at least one of the words: schistosoma schistosome schistosomiasis cercaria cercariae cercarium mansoni haematobium japonicum bilharzia bilharziasis "snail fever"

- **CNKI: Professional search**

(TI='血吸虫' OR TI='尾蚴') AND (TI='皂' OR TI='灰' OR TI='洗衣' OR TI='沐浴' OR TI='洗手' OR TI='清洁' OR TI='卫生')
